# Community knowledge, chronic morbidity and microfilaraemia following mass drug administration for lymphatic filariasis in northern Ghana: a cross-sectional study

**DOI:** 10.64898/2026.10.07.26364534

**Authors:** Cosmas Ngmaanoma Ngmembong, Millicent Captain-Esoah, Sharmila Lareef, Elijah Dakorah Angyiereyiri, Martin Ntiamoah Donkor, Sylvester Donne Dassah, Mawuli Kwamla Azameti, Grace Adzo Motey, Lawrence Adelani Adetunde, Theophilus Atio Abalori, Kwadwo Kyereme Frempong, Chrysantus Kubio, Victor Asoala, Daniel Adjei Boakye, Samuel Kweku Dadzie, Felicia Owusu-Antwi, Michael Rockson Adjei, Martin Peter Grobusch, Didier Bakajika, Frank John Lule, Elizabeth Juma, Fiona Braka, Dorothy Achu

## Abstract

**Objectives:** To assess microfilaraemia, chronic lymphatic filariasis (LF)-related morbidity, and community knowledge and self-reported preventive behaviours in a rural community in northern Ghana following prolonged mass drug administration (MDA).

**Design:** Community-based cross-sectional study.

**Setting:** Birifor community, Lawra Municipality, Upper West Region, northern Ghana, from October 2024 to January 2025.

**Participants:** A total of 261 community members aged 10 years and above were selected using random sampling and participated in the study.

**Outcome measures:** Outcome measures included microfilaraemia detected by night blood microscopy, chronic LF-related morbidity identified through clinical examination, and community knowledge and self-reported preventive behaviours assessed using a structured questionnaire.

**Results:** No microfilariae were detected (0/261; 0%). Chronic lymphoedema was identified in five individuals (1.9%), all aged over 40 years. Awareness of LF was high (95.8%), yet only 39.5% of participants demonstrated good community knowledge and self-reported preventive behaviours. Misconceptions regarding transmission, particularly beliefs that LF is hereditary or caused by spiritual factors, were common. MDA participation was high (93.1%). All five individuals with chronic lymphoedema reported loss of income, and three (60%) reported additional household income loss due to caregiving responsibilities.

**Conclusions:** No microfilaraemia was detected, suggesting very low transmission in the study area, although residual infection cannot be excluded. Chronic LF-related morbidity and gaps in community knowledge persist despite high MDA participation. Sustained surveillance using more sensitive diagnostic approaches, together with morbidity management and disability prevention, community education and support for affected households, will be important to sustain LF elimination efforts.

**Article Summary:** This community-based cross-sectional study assessed microfilaraemia, chronic lymphatic filariasis-related morbidity, and community knowledge and self-reported preventive behaviours in a rural community in northern Ghana following prolonged mass drug administration. No microfilariae were detected, suggesting very low transmission, although residual infection could not be excluded. Chronic lymphoedema persisted among a small number of older community members, and important misconceptions regarding disease transmission and prevention remained common despite high awareness and MDA participation. These findings highlight the need for continued surveillance, strengthened morbidity management and disability prevention services, and targeted community education to sustain lymphatic filariasis elimination efforts.

**Strengths and limitations of this study:**

- This study combined parasitological assessment, clinical evaluation of chronic lymphatic filariasis-related morbidity, and assessment of community knowledge and self-reported preventive behaviours within a single community-based survey.
- The study was conducted in a post-mass drug administration setting, providing evidence relevant to lymphatic filariasis elimination and post-elimination surveillance programmes.
- Use of a participant flow diagram and complete-data analysis improved transparency and reporting of participant recruitment and inclusion.
- The sample size was below the calculated minimum required for prevalence estimation, limiting the precision of prevalence estimates and the statistical power for multivariable analyses.
- Parasitological assessment relied on night blood microscopy because circulating filarial antigen detection and molecular diagnostic methods were not available during fieldwork, limiting detection of low-level residual infection.

## Introduction

Lymphatic filariasis (LF) is a mosquito-borne neglected tropical disease that can cause progressive lymphatic dysfunction, lymphoedema, hydrocele, disability and long-term psychosocial and economic consequences for affected individuals and households [1–3]. The Global Programme to Eliminate Lymphatic Filariasis (GPELF), launched in 2000, is built on two pillars: (i) annual mass drug administration (MDA) to interrupt transmission, and (ii) morbidity management and disability prevention (MMDP) to improve the quality of life of affected individuals [4–6]. As of 2024, more than 19 countries have been validated by the World Health Organization (WHO) as having eliminated LF as a public health problem, reflecting substantial reductions in transmission across many endemic settings. [5].

Despite these gains, chronic morbidity and residual transmission remain challenges in some endemic areas. Individuals may continue to experience lymphoedema or hydrocele despite reductions in infection prevalence, and geographic heterogeneity in transmission and morbidity can persist [6–11]. Community perceptions, treatment uptake, adherence to MDA, and preventive practices also influence transmission dynamics and programme effectiveness, particularly in near-elimination settings

Following the launch of GPELF, Ghana implemented annual MDA with ivermectin and albendazole across endemic districts. Many districts subsequently achieved epidemiological thresholds required to stop community-wide treatment and are now in the post-MDA surveillance phase. Nevertheless, persistent antigen positivity and reports of chronic LF-related morbidity in some northern districts highlight the need for continued surveillance and community-based assessments [8,12,13]. Studies from northern Ghana have reported higher levels of infection and morbidity than in southern regions, and persistent misconceptions about LF transmission and prevention remain a challenge [8,14,15].

Antigen detection assays and molecular methods provide greater sensitivity than microscopy for detecting low-level infections, but logistical and operational constraints often limit their routine use in endemic settings [16,17]. At the same time, long-term morbidity management remains under-prioritised despite evidence that simple limb-care practices reduce acute inflammatory episodes and slow lymphoedema progression [6,18,19].

We hypothesised that, despite prolonged MDA and the expected reduction in microfilaraemia, chronic LF-related morbidity and misconceptions regarding disease transmission and prevention would persist within the community. We therefore conducted a community-based study in Birifor, northern Ghana, to assess: (i) microfilaraemia using night blood microscopy; (ii) chronic LF-related morbidity through clinical examination; and (iii) community knowledge and self-reported preventive behaviours related to LF. We also examined associations between these outcomes and selected demographic and socioeconomic characteristics. The findings provide evidence to support surveillance, MMDP programmes, and community engagement strategies in districts approaching elimination.

## Methods

### Study Design and Period

A community-based cross-sectional survey was conducted between 15 October 2024 and 20 January 2025 to assess (i) the presence of circulating Wuchereria bancrofti microfilariae (microfilaraemia); (ii) chronic LF-related morbidity, including lymphoedema and hydrocele; and (iii) community knowledge and self-reported preventive behaviours regarding LF transmission, prevention, treatment, and morbidity management. All field procedures were conducted in accordance with WHO guidelines for LF monitoring and epidemiological assessment, as well as MMDP [19,20].

### Study Site

Birifor is a rural community located in the Lawra Municipality of the Upper West Region of Ghana (10°30’02.3’N, 002°51’43.8’W). The community has an estimated population of approximately 3,500 residents and is situated about 2 km east of Babile and 2 km west of the Black Volta River, which forms part of the international boundary between Ghana and Burkina Faso (Fig. 1). The majority of residents are subsistence farmers engaged in mixed crop cultivation and livestock rearing.

**Fig 1.**
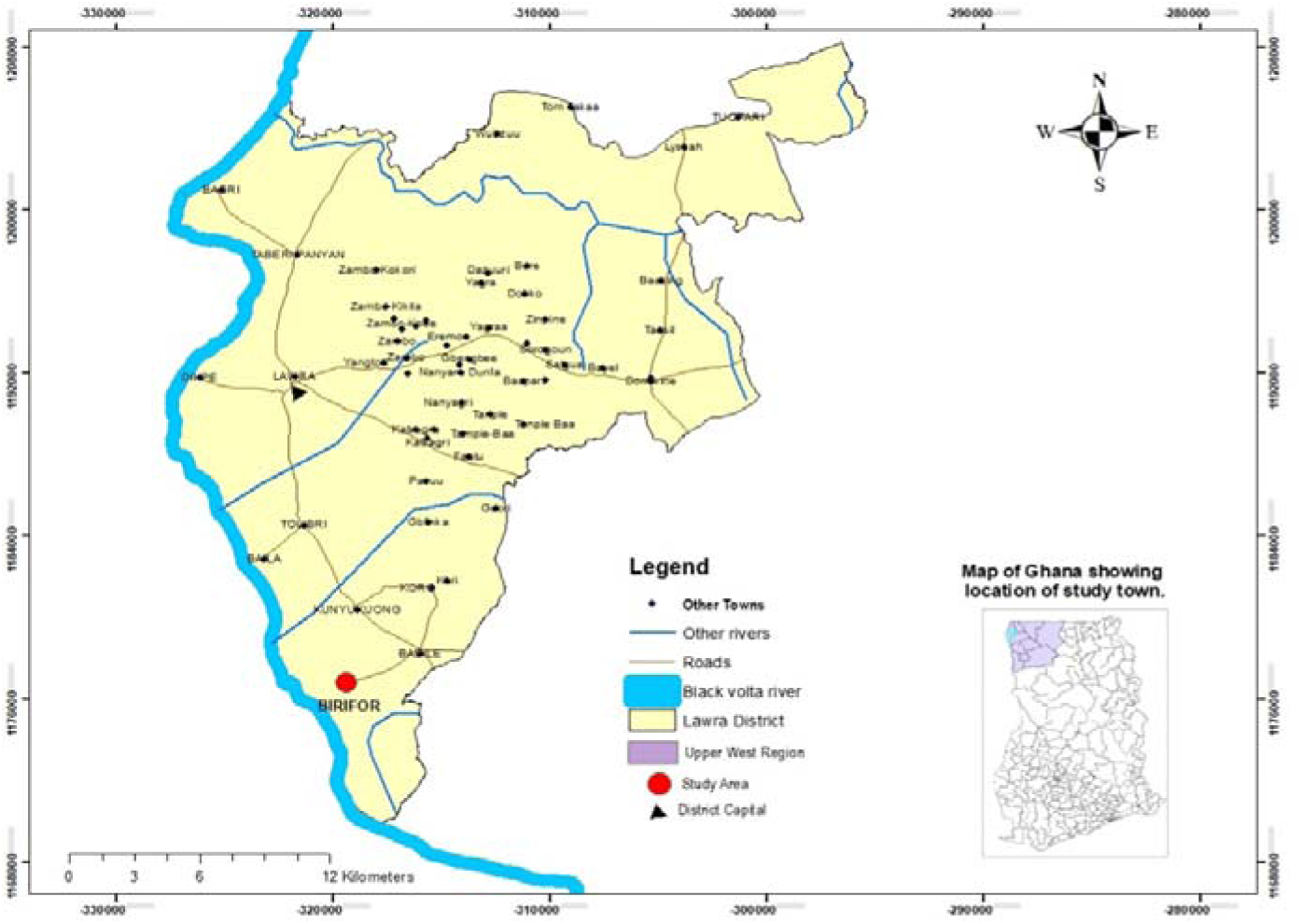
Map of Lawra Municipality showing the location of Birifor community (study area) in the Upper West Region of northern Ghana. The map was created by the authors using ArcGIS with district administrative boundary shapefiles from the HDX Ghana Administrative Boundaries dataset (CC BY-IGO).

The area has historically been endemic for LF and has received multiple rounds of annual MDA under Ghana’s national LF elimination programme. Despite substantial progress towards elimination, concerns remain regarding residual infection, chronic LF-related morbidity, and community awareness. Birifor was therefore selected as the study site for a comprehensive assessment of LF in a near-elimination setting.

### Eligibility criteria

Individuals aged 10 years and above who had resided in Birifor for at least five years were eligible for inclusion. The residency requirement was intended to ensure adequate exposure to local LF transmission patterns, control interventions, and community health education activities.

Participants were excluded if they had taken antifilarial medications (diethylcarbamazine, ivermectin, or albendazole) within the preceding three months, declined to provide written informed consent (or assent for minors), or were unwilling to provide night blood samples, undergo clinical examination, or complete the study questionnaire.

The most recent MDA round in the study area was conducted in May 2024, with reported treatment coverage exceeding 80%. The community survey commenced five months after this MDA round. The exclusion of individuals who had received antifilarial treatment within the preceding three months was intended to minimise the potential for transient suppression of microfilaraemia, which could reduce the sensitivity of microscopy-based detection.

### Sample Size and Sampling Strategy

The sample size was determined using Cochran’s formula, assuming an expected prevalence of 50%, a 95% confidence level (Z = 1.96), and a 5% margin of error. This yielded an initial sample size of 384 participants. After applying a finite population correction for the estimated population of 3,500, the minimum required sample size was 346. Due to logistical and operational constraints, 261 participants were enrolled. As this was below the calculated minimum sample size, the precision of the prevalence estimates was reduced and is acknowledged as a study limitation. Furthermore, the sample size calculation was based on prevalence estimation and was not designed to provide adequate statistical power for multivariable regression analyses.

The sampling frame was derived from the 2021 Population and Housing Census, which enumerated approximately 3,500 residents in the community [21]. Eligible individuals were assigned unique identification numbers and selected by simple random sampling using a computer-generated random number generator. A total of 286 individuals were approached for participation. Of these, 261 consented and were enrolled in the study. Ten individuals were excluded because they did not meet the eligibility criteria, primarily owing to residence in the community for less than five years, and 15 declined to participate. Non-responders and ineligible individuals were not replaced, and recruitment ceased after 261 participants had been enrolled because of logistical and operational constraints (Fig. 2).

**Fig 2.**
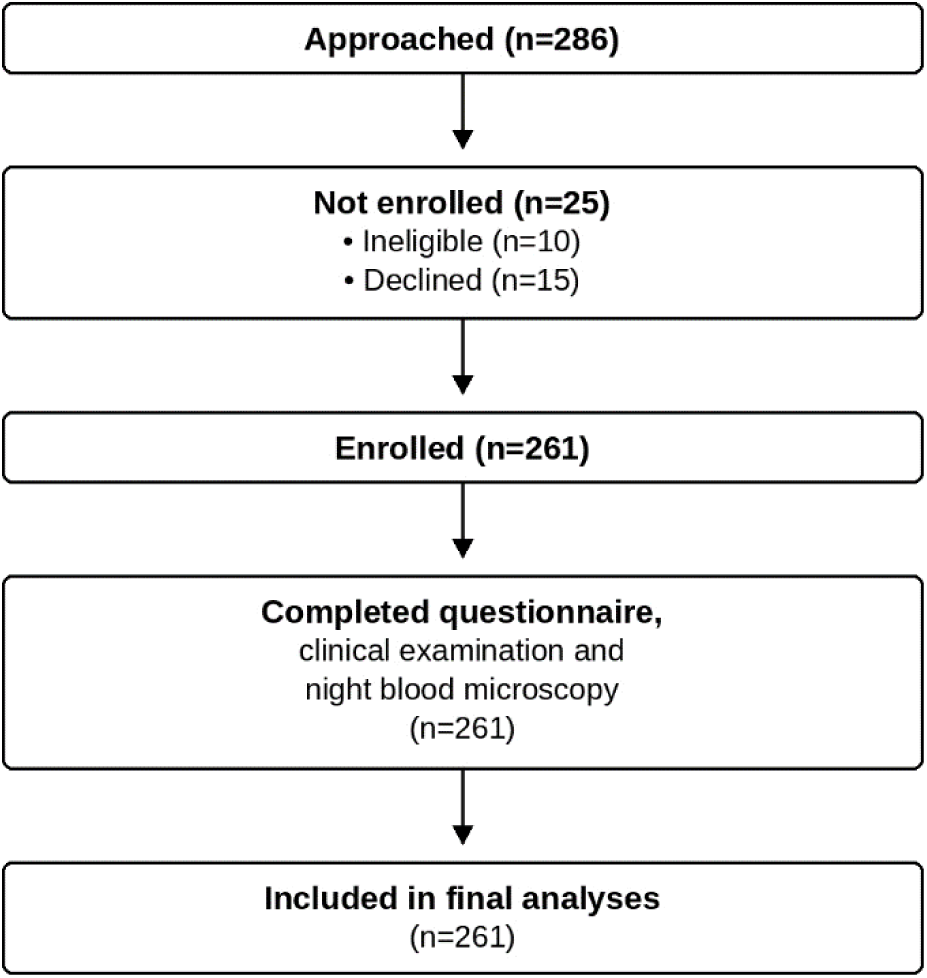
Flow diagram showing participant recruitment, enrolment, and inclusion in the final analyses.

### Data Collection Procedures

Data were collected between 15 October 2024 and 20 January 2025 by trained field workers using standardized data collection procedures. A structured questionnaire was administered to collect socio-demographic information and selected indicators of community knowledge and self reported preventive behaviours related to LF, including knowledge of disease transmission, prevention, treatment, morbidity management, and participation in MDA. The questionnaire used for data collection is provided as online supplementary file 1. Although the questionnaire included items on community knowledge, perceptions, and self-reported preventive behaviours, it was not designed as a comprehensive knowledge, attitudes, and practices (KAP) survey. Consequently, the indicators reported in this study represent a focused assessment of selected knowledge and behavioural domains rather than a comprehensive evaluation of all behavioural and perceptual dimensions related to LF.

Clinical examinations were conducted by trained clinicians to identify signs of chronic LF-related morbidity, including lymphoedema and hydrocele. Chronic LF-related morbidity was defined as the presence of lymphoedema of the limbs and/or hydrocele in male participants, as determined by clinical examination. Night blood samples were collected from eligible participants to detect circulating *Wuchereria bancrofti* microfilariae, as described below.

### Parasitological assessment

Night blood samples were collected from eligible participants by certified medical laboratory scientists between 22:00 and 02:00 hours to account for the nocturnal periodicity of *Wuchereria bancrofti* microfilariae. Both capillary and venous blood samples were collected. Thin blood smears were prepared from capillary blood, while thick smears were prepared from venous blood samples. All smears were stained using standard laboratory procedures and examined microscopically for the presence of *Wuchereria bancrofti* microfilariae by experienced medical laboratory scientists.

Only night blood microscopy was performed because circulating filarial antigen detection assays and molecular diagnostic methods were not available during fieldwork. Consequently, circulating filarial antigenaemia and molecular evidence of residual infection could not be assessed.

### Data management and analysis

Data collected using Kobo Collect were exported into Microsoft Excel and subsequently analysed using Stata version 17 (StataCorp LLC, College Station, TX, USA). Data completeness was assessed before analysis. No missing observations were identified for variables included in the descriptive or multivariable analyses, and all enrolled participants (n = 261) were included in the final analyses.

Descriptive statistics were used to summarise participants’ socio-demographic characteristics and questionnaire responses. Associations between categorical variables and the composite community knowledge and self-reported preventive behaviour outcome were assessed using Pearson’s Chi-square test or Fisher’s exact test, as appropriate.

Community knowledge and self-reported preventive behaviours were summarised using a classification adapted from previous LF studies conducted in Ghana and similar endemic settings [9,22]. Four domains were assessed: knowledge of disease transmission, prevention, signs and symptoms, and awareness of MDA. Each correct response was assigned a score of 1, and each incorrect response a score of 0. The four domains were equally weighted, yielding a total score ranging from 0 to 4. Participants who scored 4 were classified as having good community knowledge and self-reported preventive behaviours, whereas those scoring less than 4 were classified as having limited community knowledge and self-reported preventive behaviours. This approach distinguished participants with complete knowledge across all assessed domains from those with knowledge gaps in one or more domains.

Logistic regression analysis was performed to examine associations between the composite community knowledge and self-reported preventive behaviour outcome and selected socio-demographic characteristics. Crude odds ratios (cORs) with 95% confidence intervals (CIs) were estimated using bivariable logistic regression. Variables considered epidemiologically relevant based on prior literature, including age, marital status, education, and occupation, were entered into the multivariable logistic regression model, irrespective of their statistical significance in the bivariable analysis, to adjust for potential confounding. Adjusted odds ratios (aORs) with 95% CIs were then estimated from the multivariable model. Statistical significance was set at p < 0.05.

Model diagnostics were performed following multivariable logistic regression. Multicollinearity was assessed using variance inflation factors (VIFs). Model calibration was assessed using the Hosmer-Lemeshow goodness-of-fit test, model discrimination was assessed using the area under the receiver operating characteristic (ROC) curve, and model explanatory power was assessed using McFadden’s pseudo-R².

### Ethical Considerations

Ethical approval for the study was obtained from the Navrongo Health Research Centre Institutional Review Board (NHRCIRB; Reference No. NHRCIRB658; approved on 8 October 2024). Additional administrative permission was obtained from the Lawra Municipal Health Directorate and relevant community leaders. Written informed consent was obtained from all adult participants. For participants younger than 18 years, written assent was obtained in addition to written informed consent from a parent or legal guardian. Participation was voluntary, and the confidentiality of all participant information was strictly maintained.

Participants identified with chronic lymphoedema were enrolled in a community care programme, where they received education on lymphoedema self-management together with medical supplies, including bandages to support self-care. An assigned healthcare worker conducted monthly follow-up visits to monitor progress, reinforce recommended care practices, and provide ongoing support. Following completion of the study, the findings and recommendations were shared with community members, community leaders, and local health authorities to support community-led action and inform ongoing lymphatic filariasis elimination efforts.

### Patient and Public Involvement

Patients and/or the public were not involved in the design, conduct, reporting, or dissemination plans of this research. Following completion of the study, findings and recommendations were shared with community members, community leaders, and local health authorities to support community-led action and ongoing lymphatic filariasis elimination efforts.

### Reporting Guideline

This study is reported in accordance with the Strengthening the Reporting of Observational Studies in Epidemiology (STROBE) guideline for cross-sectional studies. The completed STROBE checklist is provided as a supplementary file.

## Results

### Sociodemographic Characteristics, Knowledge, and Preventive Behaviours

A total of 261 participants were included in the analysis. Overall, 103 (39.5%) participants were classified as having good knowledge and preventive behaviours related to lymphatic filariasis, while 158 (60.5%) had limited knowledge and preventive behaviours. Participants aged 10–30 years constituted the largest age group (61.7%), followed by those aged 31–60 years (30.3%) and those aged >60 years (8.0%). The proportion classified as having good knowledge and preventive behaviours increased with age, from 34.2% among participants aged 10–30 years to 45.6% among those aged 31–60 years and 57.1% among those aged >60 years. This difference was statistically significant (p=0.010).

Males constituted 53.7% of the participants and females 46.3%. Good knowledge and preventive behaviours were reported among 40.7% of males and 38.0% of females, with no statistically significant difference by sex (p=0.657). Regarding marital status, 47.6% of participants had never married, 43.3% were married, and 9.2% were widowed or divorced. The proportion with good knowledge and preventive behaviours was lower among participants who had never married (29.8%) than among married participants (48.7%) and those who were widowed or divorced (45.8%), with a statistically significant association observed between marital status and the classification (p=0.037).

Educational attainment was also significantly associated with knowledge and preventive behaviours (p=0.020). Participants with tertiary education had the highest proportion classified as having good knowledge and preventive behaviours (66.7%), compared with 41.2% among those with no formal education, 30.4% among those with primary education, and 41.0% among those with secondary education.

Farmers constituted the largest occupational group (46.0%), followed by students (27.2%), traders (17.6%), public sector workers (6.5%), and unemployed participants (2.7%). The proportion with good knowledge and preventive behaviours ranged from 29.6% among students to 64.7% among public sector workers. However, the association between occupation and knowledge and preventive behaviours was not statistically significant (p=0.086) (Table 1).

**Table 1.**
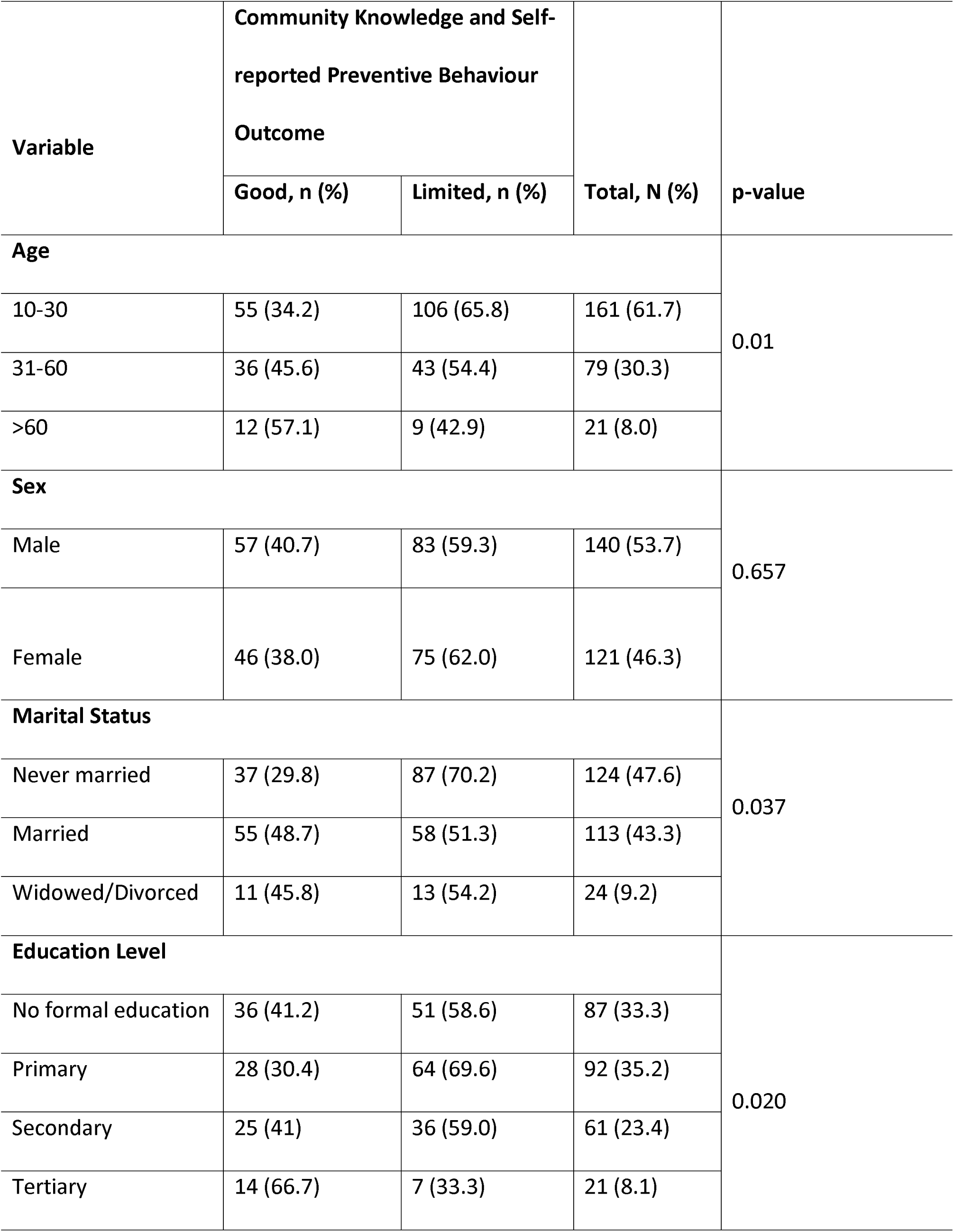
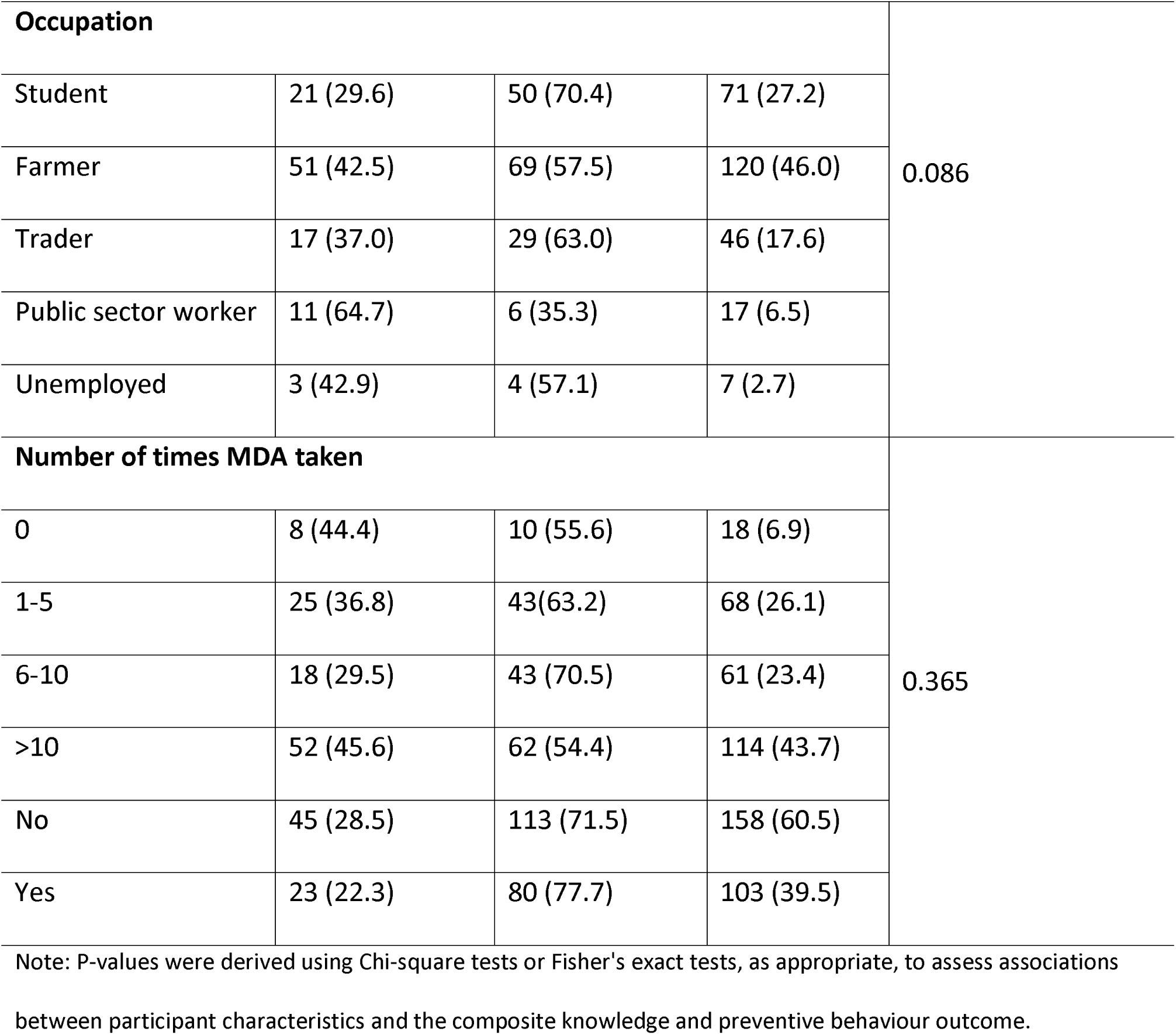
Sociodemographic characteristics and binary classification of knowledge and preventive behaviours related to lymphatic filariasis among study participants in Birifor, Ghana

| Variable | Community Knowledge and Self-reported Preventive Behaviour Outcome |  | Total, N (%) | p-value |
| --- | --- | --- | --- | --- |
|  | Good, n (%) | Limited, n (%) |  |  |
| Age |  |  |  | 0.01 |
| 10-30 | 55 (34.2) | 106 (65.8) | 161 (61.7) |  |
| 31-60 | 36 (45.6) | 43 (54.4) | 79 (30.3) |  |
| >60 | 12 (57.1) | 9 (42.9) | 21 (8.0) |  |
| Sex |  |  |  | 0.657 |
| Male | 57 (40.7) | 83 (59.3) | 140 (53.7) |  |
| Female | 46 (38.0) | 75 (62.0) | 121 (46.3) |  |
| Marital Status |  |  |  | 0.037 |
| Never married | 37 (29.8) | 87 (70.2) | 124 (47.6) |  |
| Married | 55 (48.7) | 58 (51.3) | 113 (43.3) |  |
| Widowed/Divorced | 11 (45.8) | 13 (54.2) | 24 (9.2) |  |
| Education Level |  |  |  | 0.020 |
| No formal education | 36 (41.2) | 51 (58.6) | 87 (33.3) |  |
| Primary | 28 (30.4) | 64 (69.6) | 92 (35.2) |  |
| Secondary | 25 (41) | 36 (59.0) | 61 (23.4) |  |
| Tertiary | 14 (66.7) | 7 (33.3) | 21 (8.1) |  |

| Occupation |  |  |  | 0.086 |
| --- | --- | --- | --- | --- |
| Student | 21 (29.6) | 50 (70.4) | 71 (27.2) |  |
| Farmer | 51 (42.5) | 69 (57.5) | 120 (46.0) |  |
| Trader | 17 (37.0) | 29 (63.0) | 46 (17.6) |  |
| Public sector worker | 11 (64.7) | 6 (35.3) | 17 (6.5) |  |
| Unemployed | 3 (42.9) | 4 (57.1) | 7 (2.7) |  |
| Number of times MDA taken |  |  |  | 0.365 |
| 0 | 8 (44.4) | 10 (55.6) | 18 (6.9) |  |
| 1-5 | 25 (36.8) | 43 (63.2) | 68 (26.1) |  |
| 6-10 | 18 (29.5) | 43 (70.5) | 61 (23.4) |  |
| >10 | 52 (45.6) | 62 (54.4) | 114 (43.7) |  |
| No | 45 (28.5) | 113 (71.5) | 158 (60.5) |  |
| Yes | 23 (22.3) | 80 (77.7) | 103 (39.5) |  |

Note: P-values were derived using Chi-square tests or Fisher's exact tests, as appropriate, to assess associations between participant characteristics and the composite knowledge and preventive behaviour outcome.

### Community knowledge and self reported preventive behaviours regarding lymphatic filariasis transmission, clinical manifestation and prevention

Overall awareness of LF was high, with 250 of 261 participants (95.8%) reporting having heard of the disease. Despite this high level of awareness, important misconceptions about disease transmission persisted. Only 123 participants (47.1%) correctly identified mosquito bites as the mode of transmission, whereas others attributed LF to person-to-person contact (28.0%), spiritual causes (14.9%), or heredity (10.0%) (Fig. 3A).

**Fig 3.**
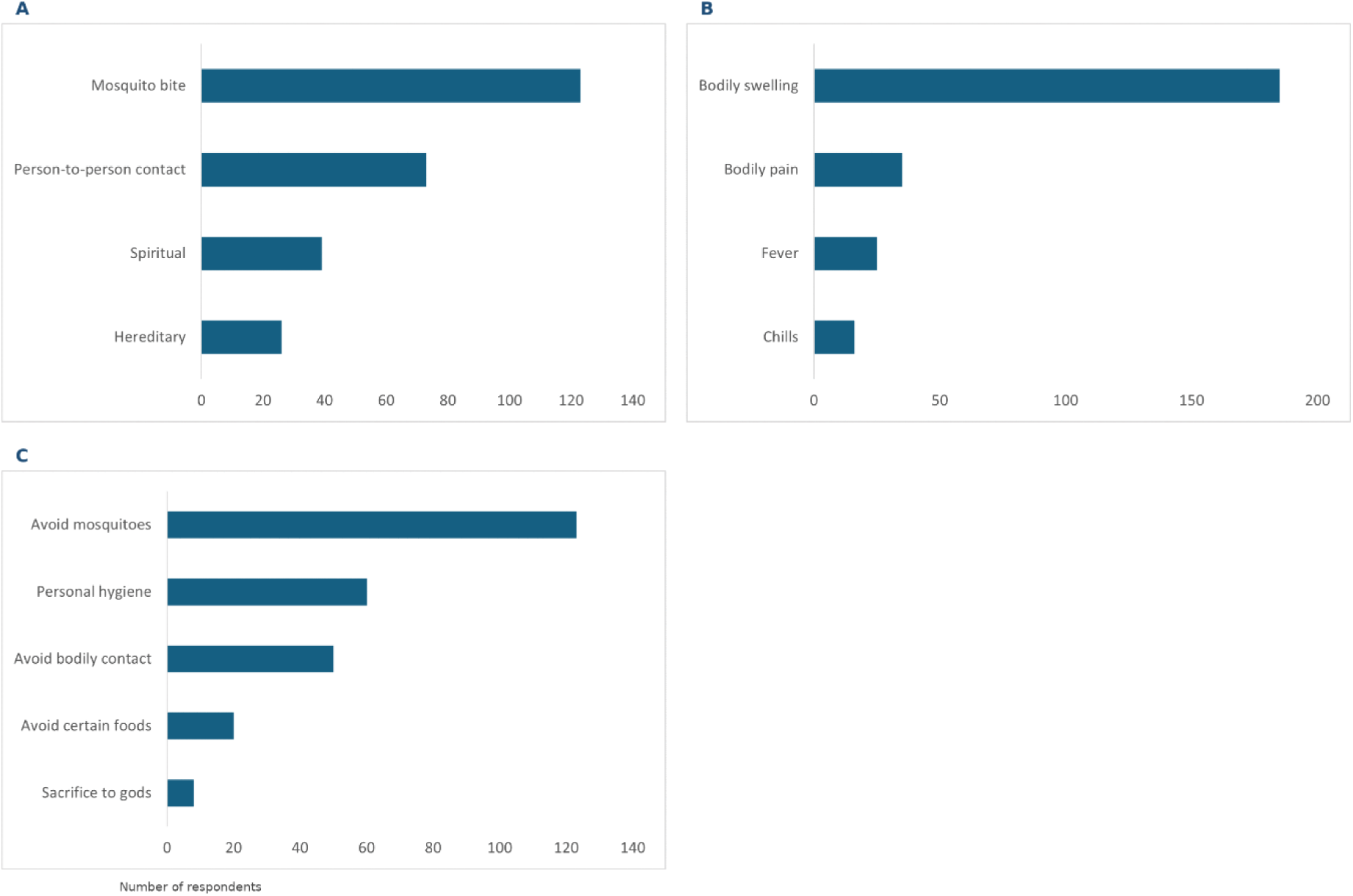
Community knowledge of lymphatic filariasis transmission, clinical manifestations, and prevention. in Birifor, Ghana. Panel A: Community knowledge of lymphatic filariasis transmission, Panel B: Community knowledge of clinical manifestations of lymphatic filariasis, and Panel C: Community knowledge of lymphatic filariasis prevention. Values represent mutually exclusive participant responses (N=261).

Most participants (185 of 261; 70.9%) correctly recognized limb or genital swelling as a key clinical manifestation of LF (Fig. 3B).

Regarding prevention, 123 participants (47.1%) identified protection against mosquito bites as a preventive measure, while 60 (23.0%) mentioned personal hygiene practices. Notably, 50 participants (19.2%) cited avoiding bodily contact with affected individuals as a preventive measure, reflecting persistent misconceptions about LF transmission (Fig. 3C).

### Participation in MDA

Awareness of the national MDA programme was high, with 250 of 261 participants (95.8%) reporting prior knowledge of the intervention. A total of 243 participants (93.1%) reported having ever taken antifilarial medications during MDA campaigns, and nearly half (114 of 261; 43.7%) reported participating in more than 10 rounds of MDA. Most respondents (158 of 261; 60.5%) reported no adverse effects following treatment (Fig. 4).

**Fig 4:**
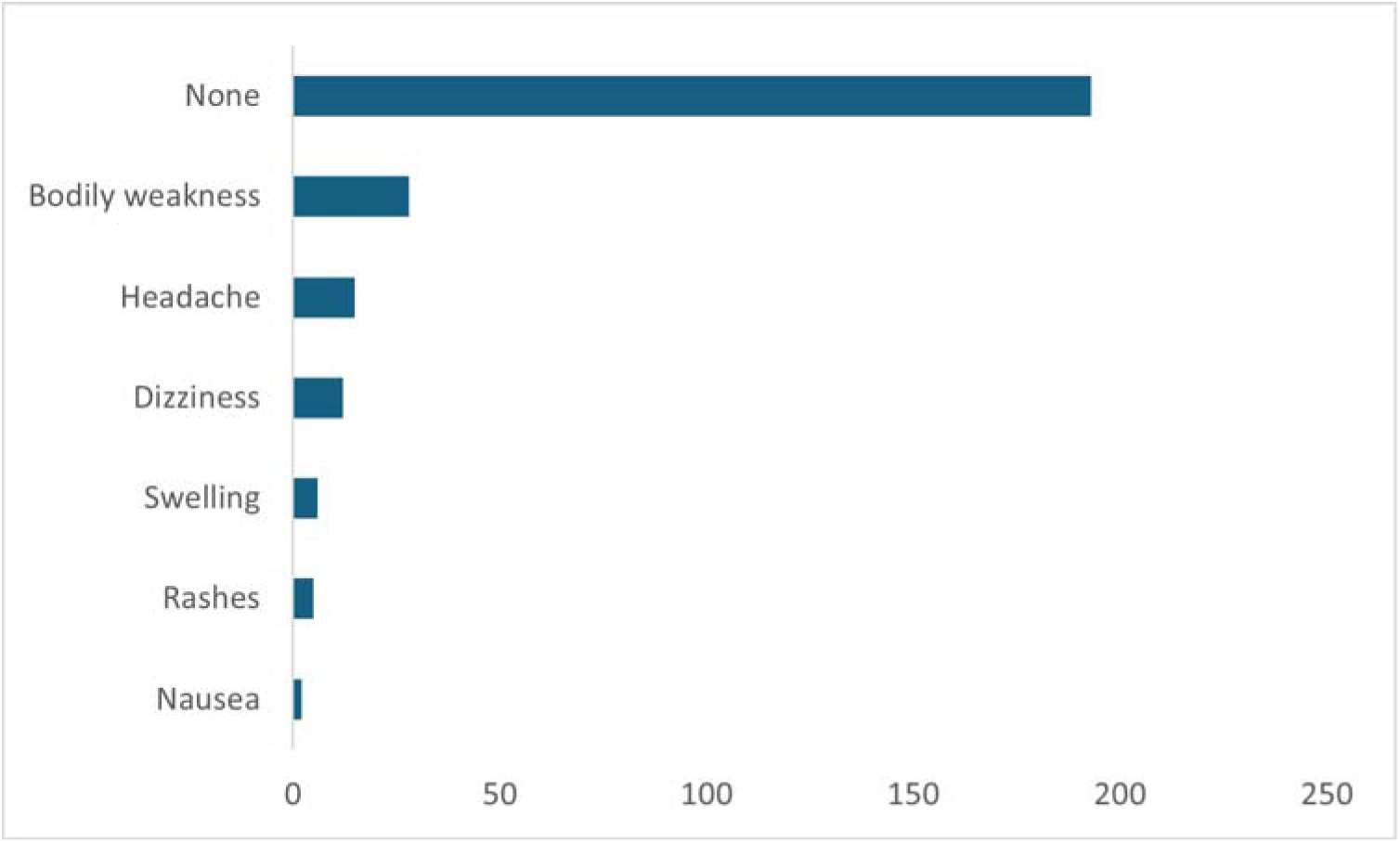
Adverse effects reported after mass drug administration in Birifor, Ghana. Participants could report more than one adverse effect; therefore, the total number of responses exceeds the number of participants (N = 261).

### Prevalence of Microfilariae and Chronic Morbidity

Microscopic examination of night blood smears did not detect microfilariae in any participant (0 of 261), corresponding to an estimated prevalence of 0% (95% CI: 0–1.1%). Chronic LF-related morbidity was identified in five participants, corresponding to a prevalence of 1.9% (5 of 261). Of these, three cases involved lower limb lymphoedema (all male), while two involved upper limb lymphoedema (one male and one female). All affected individuals were aged above 40 years. No cases of hydrocele were identified among the male participants.

### Impact of Chronic Morbidity on Daily Life

All participants with chronic LF manifestations (5 of 5; 100%) reported loss of income due to functional limitations associated with the condition. In addition, three participants (60%) reported that other household members had lost income due to caregiving responsibilities.

### Associations Between Sociodemographic Characteristics and Community Knowledge and Self-reported Preventive Behaviours

At the bivariate level, several sociodemographic characteristics were significantly associated with good community knowledge and self-reported preventive behaviours related to LF. These included age greater than 60 years (cOR = 2.57, 95% CI: 1.02–6.47; p = 0.045), being married (cOR = 2.23, 95% CI: 1.31–3.80; p = 0.003), having tertiary education (cOR = 2.83, 95% CI: 1.04– 7.72; p = 0.042), and employment in the public sector (cOR = 4.37, 95% CI: 1.43–13.35; p = 0.010). Although the overall association between occupation and the outcome was not statistically significant (Fisher’s exact test, p = 0.086), public sector workers had significantly higher odds of good community knowledge and self-reported preventive behaviours than students in the unadjusted analysis. However, none of the sociodemographic characteristics remained statistically significant after adjustment for potential confounders in the multivariable logistic regression model (Table 2).

**Table 2.**
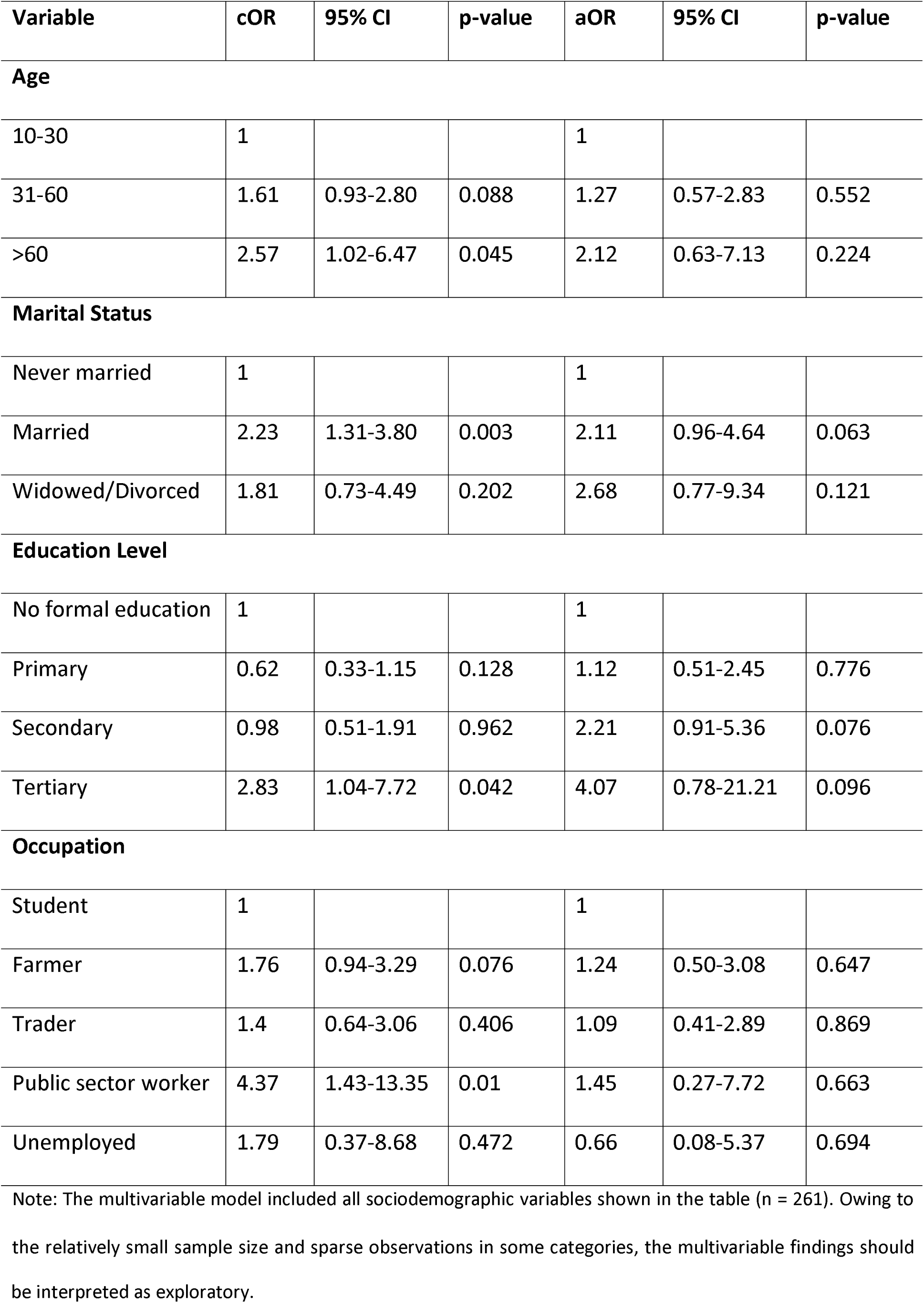
Factors associated with good community knowledge and self-reported preventive behaviours related to lymphatic filariasis among community members in Birifor District, Ghana

| Variable | cOR | 95% CI | p-value | aOR | 95% CI | p-value |
| --- | --- | --- | --- | --- | --- | --- |
| **Age** |  |  |  |  |  |  |
| 10-30 | 1 |  |  | 1 |  |  |
| 31-60 | 1.61 | 0.93-2.80 | 0.088 | 1.27 | 0.57-2.83 | 0.552 |
| >60 | 2.57 | 1.02-6.47 | 0.045 | 2.12 | 0.63-7.13 | 0.224 |
| **Marital Status** |  |  |  |  |  |  |
| Never married | 1 |  |  | 1 |  |  |
| Married | 2.23 | 1.31-3.80 | 0.003 | 2.11 | 0.96-4.64 | 0.063 |
| Widowed/Divorced | 1.81 | 0.73-4.49 | 0.202 | 2.68 | 0.77-9.34 | 0.121 |
| **Education Level** |  |  |  |  |  |  |
| No formal education | 1 |  |  | 1 |  |  |
| Primary | 0.62 | 0.33-1.15 | 0.128 | 1.12 | 0.51-2.45 | 0.776 |
| Secondary | 0.98 | 0.51-1.91 | 0.962 | 2.21 | 0.91-5.36 | 0.076 |
| Tertiary | 2.83 | 1.04-7.72 | 0.042 | 4.07 | 0.78-21.21 | 0.096 |
| **Occupation** |  |  |  |  |  |  |
| Student | 1 |  |  | 1 |  |  |
| Farmer | 1.76 | 0.94-3.29 | 0.076 | 1.24 | 0.50-3.08 | 0.647 |
| Trader | 1.4 | 0.64-3.06 | 0.406 | 1.09 | 0.41-2.89 | 0.869 |
| Public sector worker | 4.37 | 1.43-13.35 | 0.01 | 1.45 | 0.27-7.72 | 0.663 |
| Unemployed | 1.79 | 0.37-8.68 | 0.472 | 0.66 | 0.08-5.37 | 0.694 |

Note: The multivariable model included all sociodemographic variables shown in the table (n = 261). Owing to the relatively small sample size and sparse observations in some categories, the multivariable findings should
be interpreted as exploratory.

Model diagnostics indicated no evidence of problematic multicollinearity among the predictor variables, with all variance inflation factors below the commonly accepted threshold of 5. The Hosmer–Lemeshow goodness-of-fit test indicated adequate model fit (χ² = 7.75, df = 8, p = 0.458). The model demonstrated an area under the receiver operating characteristic (ROC) curve of 0.66 and a McFadden pseudo-R² of 0.06, indicating modest discriminatory ability and explanatory power.

## Discussion

This study assessed the epidemiological status of LF, together with community knowledge and self-reported preventive behaviours, in a rural Ghanaian setting nearly two decades after implementation of elimination strategies. No microfilariae were detected in the study population, suggesting that transmission has been substantially reduced. However, these findings should be interpreted cautiously because the study did not achieve the calculated sample size, and microscopy has limited sensitivity for detecting low-density infections in near-elimination settings. Consequently, residual infection cannot be excluded. These findings are consistent with reports from Ghana and other endemic settings demonstrating the long-term impact of sustained MDA programmes on reducing LF transmission [12,15,22]. Despite the absence of detectable microfilaraemia, chronic LF-related morbidity persisted (1.9%) highlighting that interruption of transmission does not necessarily eliminate the existing disease burden.

Despite high awareness of LF (95.8%), only 39.5% of participants were classified as having good community knowledge and self-reported preventive behaviour indicators. Misconceptions regarding transmission, particularly beliefs attributing LF to heredity, person-to-person contact, or spiritual causes, remained prevalent. These findings are consistent with previous studies documenting persistent misconceptions and cultural interpretations of LF in endemic communities [9,23–25]. Although nearly half of the participants correctly identified mosquito bites as the mode of transmission, the findings indicate persistent gaps in community knowledge regarding the disease. Nevertheless, the relatively high awareness of mosquito-borne transmission provides an opportunity to strengthen behaviour change communication and reinforce vector control measures, including the use of insecticide-treated nets, as part of efforts to sustain LF elimination [26–28].

The high reported uptake of MDA (93.1%) reflects strong programme reach and community engagement. This is consistent with evidence from Ghana and other endemic settings demonstrating that repeated rounds of MDA substantially reduce LF transmission and infection prevalence [12,15]. The low frequency of reported adverse effects further supports the safety and acceptability of antifilarial medicines, consistent with findings from other endemic settings where most participants reported minimal or no side effects [12,24,29]. Nevertheless, maintaining high MDA coverage until elimination targets are achieved remains essential, as even small groups of untreated individuals may sustain residual transmission and delay interruption of transmission [9,11,30–32].

The persistence of chronic LF-related morbidity observed in this study is consistent with reports from other endemic regions transitioning towards elimination, where transmission declines but established disease remains [5,6,11,33]. Chronic manifestations such as lymphoedema result from irreversible lymphatic damage and may persist even after parasite clearance. The prevalence of chronic LF-related morbidity (1.9%) was higher than that reported in some post-MDA settings [34,35], possibly reflecting historically higher transmission intensity and periods of suboptimal MDA coverage. Importantly, chronic LF-related morbidity was associated with a considerable socioeconomic burden, with all affected individuals reporting loss of income and additional household-level economic strain. Although these findings were based on only five participants with chronic morbidity, similar socioeconomic consequences, including stigma, reduced productivity, and increased household vulnerability, have been documented in other endemic settings [36–39].

These findings highlight the need to strengthen morbidity management and disability prevention (MMDP) as a core component of LF elimination strategies. Evidence indicates that simple limb-care practices can reduce acute inflammatory episodes and slow disease progression [40], yet such interventions remain under-prioritised in many endemic settings [33]. Integrating MMDP into routine primary health care systems is essential to improve access to care and ensure long-term sustainability. Furthermore, the coexistence of no detectable microfilaraemia, residual morbidity, and persistent knowledge gaps underscores the need for a dual-track strategy in near-elimination settings that sustains post-MDA surveillance while addressing the long-term needs of people living with chronic LF. This should include embedding LF indicators within routine health information systems, strengthening community health worker engagement, and implementing targeted behaviour change communication interventions. Similar integrated approaches have been shown to strengthen programme ownership and improve the long-term impact of neglected tropical disease programmes [22].

This study has several strengths. The use of population-based random sampling based on an updated census enhanced the representativeness of the study population. The integration of parasitological, clinical, and community knowledge and self-reported preventive behaviour data enabled a comprehensive assessment of infection, chronic morbidity, and community knowledge in a near-elimination setting, providing a more holistic understanding of the epidemiology of lymphatic filariasis than would have been achieved using a single-domain approach. Standardised night blood microscopy performed by trained personnel further enhanced the reliability of the parasitological findings. Conducting the study in a community approaching elimination provided valuable insights into the challenges that persist despite prolonged implementation of elimination strategies, including residual morbidity and community knowledge gaps. Finally, complete data were available for all enrolled participants, eliminating the need for imputation or exclusion of observations from the analyses and strengthening the robustness of the study findings.

Despite these strengths, several limitations should be considered. First, the cross-sectional design precludes causal inference and does not capture temporal or seasonal variation in LF transmission. Second, the study did not achieve the calculated minimum sample size, enrolling 261 participants compared with the target of 346. This reduced the precision of prevalence estimates and may have introduced selection bias if enrolled participants differed systematically from eligible individuals who were not included. Information on MDA participation and adverse effects was self-reported and may therefore be subject to recall bias. In addition, reliance on microscopy alone may have reduced the sensitivity for detecting low-density infections, particularly in near-elimination settings. Consequently, residual infection and ongoing transmission cannot be excluded, and the absence of detectable microfilaraemia should not be interpreted as confirmation of elimination without complementary evidence from antigen-detection assays or molecular surveillance approaches [17,41]. Third, the absence of WHO-recommended lymphoedema staging and information on disease duration limited the characterisation of chronic morbidity and its comparability with other studies. Fourth, the relatively small number of participants with chronic LF-related morbidity limited subgroup analyses and the generalisability of estimates of their socioeconomic impact. Finally, the sample size was not designed to provide adequate statistical power for multivariable regression analyses, and several adjusted odds ratios had wide confidence intervals. Consequently, the regression findings should be interpreted as exploratory and hypothesis-generating. Although model diagnostics indicated no evidence of problematic multicollinearity and acceptable overall model fit, the relatively small sample size and sparse observations in some subgroups may have reduced the precision of the adjusted estimates. Additionally, the indicators used to assess community knowledge and self-reported preventive behaviours did not capture all behavioural and perceptual dimensions relevant to LF.

Notwithstanding these limitations, the findings relating to the absence of detectable microfilaraemia, the persistence of chronic LF-related morbidity, and remaining community knowledge gaps provide valuable insights for similar rural, post-MDA settings in northern Ghana and other endemic regions where transmission has declined but morbidity persists. Future research should evaluate more sensitive diagnostic approaches, including antigen-detection assays and molecular methods, to identify residual infection [17,41]. Longitudinal studies are also needed to better understand the progression of chronic morbidity and its long-term socioeconomic consequences. In addition, implementation research should evaluate strategies for integrating morbidity management and disability prevention (MMDP), referral systems, and social support into routine health services, while assessing behaviourally informed interventions to address persistent misconceptions and strengthen community engagement.

Collectively, these findings highlight the evolving epidemiology of LF in near-elimination settings, where programme priorities are shifting from interrupting transmission to sustaining surveillance, addressing residual morbidity, strengthening community engagement, and preventing disease resurgence.

## Conclusion

This study provides evidence from a rural community in northern Ghana approaching LF elimination. Although no microfilariae were detected, the persistence of chronic LF-related morbidity and substantial gaps in community knowledge indicate that important challenges remain beyond interruption of transmission. Misconceptions regarding disease transmission and prevention persisted despite high awareness of the disease and high participation in mass drug administration. Given the study’s reliance on microscopy and the reduced sample size, residual infection and ongoing transmission cannot be definitively excluded.

To sustain elimination gains, targeted community health education should be strengthened to address persistent misconceptions and reinforce appropriate preventive behaviours. In addition, MMDP services should be integrated into routine primary health care to improve access to care and support for affected individuals. Continued surveillance using more sensitive diagnostic approaches, including circulating filarial antigen detection assays and molecular methods, where feasible, will be important for identifying residual infection and preventing disease resurgence. Collectively, these measures will help consolidate progress towards the sustainable elimination of LF as a public health problem.

## Data Availability

Data are available upon reasonable request. Deidentified participant-level data underlying the findings of this study are available from the corresponding author upon reasonable request. Access may be subject to approval from the relevant ethical and institutional authorities and will be considered in accordance with participant consent, ethical approvals and applicable data-sharing requirements.

https://zenodo.org/records/21789968

## Declarations

### Patient consent for publication

Not required

### Author contributions

Conceptualisation: CNN, MC-E, MRA, SL, EDA and CK.

Methodology: CNN, SL, MRA, MND, SDD, MKA, GAM, LAA, MC-E, KKF and CK.

Investigation: CNN, MND, SDD, MKA, GAM, LAA, TAA, MC-E, MRA, KKF and CK.

Formal analysis: CNN, MRA, SL, VA, DAB and SKD.

Validation: SL, MRA, CNN, VA, DAB and SKD.

Writing – original draft: SL, MRA and CNN.

Writing – review and editing: SL, MRA, CNN, MC-E, EDA, CK, MND, SDD, MKA, GAM, LAA, TAA, KKF, VA, DAB, SKD, FO-A, MPG, DB, FJL, EJ, FB, and DA.

Supervision: MC-E, MRA, SL, FO-A, MPG, DB, FJL, EJ, FB and DA.

All authors reviewed and approved the final manuscript and agree to be accountable for all aspects of the work.

Guarantor: Dr Sharmila Lareef accepts full responsibility for the overall content of the work, had access to the data, and controlled the decision to publish.

## Acknowledgements

The authors thank the study participants, community leaders, field workers, and the Lawra Municipal Health Directorate for their support and cooperation during the conduct of this study. We also acknowledge the contributions of the medical laboratory scientists and healthcare personnel who participated in data collection and clinical assessments.

Artificial intelligence-assisted tools (Grammarly) were used to support language refinement, editing, and manuscript formatting. The AI-assisted suggestions were reviewed, verified, and edited by the authors, who take full responsibility for the accuracy, integrity, and final content of the manuscript.

## Supplementary file

Supplementary file 1: Study Questionnaire. Structured questionnaire used to collect sociodemographic characteristics, knowledge of lymphatic filariasis, mass drug administration participation, preventive behaviours, and chronic lymphatic filariasis-related morbidity among study participants.

Supplementary file 2: STROBE checklist for cross-sectional studies.

